# Data-Driven Prediction of COVID-19 Cases in Germany for Decision Making

**DOI:** 10.1101/2021.06.21.21257586

**Authors:** Lukas Refisch, Fabian Lorenz, Torsten Riedlinger, Hannes Taubenböck, Martina Fischer, Linus Grabenhenrich, Martin Wolkewitz, Harald Binder, Clemens Kreutz

## Abstract

**Background:** The COVID-19 pandemic has led to a high interest in mathematical models describing and predicting the diverse aspects and implications of the virus outbreak. Model results represent an important part of the information base for the decision process on different administrative levels. The Robert-Koch-Institute (RKI) initiated a project whose main goal is to predict COVID-19-specific occupation of beds in intensive care units: *Steuerungs-Prognose von Intensivmedizinischen COVID-19 Kapazitäten* (SPoCK). The incidence of COVID-19 cases is a crucial predictor for this occupation.

**Methods:** We developed a model based on ordinary differential equations for the COVID-19 spread with a time-dependent infection rate described by a spline. Furthermore, the model explicitly accounts for weekday-specific reporting and adjusts for reporting delay. The model is calibrated in a purely data-driven manner by a maximum likelihood approach. Uncertainties are evaluated using the profile likelihood method. The uncertainty about the appropriate modeling assumptions can be accounted for by including and merging results of different modelling approaches.

**Results:** The model is calibrated based on incident cases on a daily basis and provides daily predictions of incident COVID-19 cases for the upcoming three weeks including uncertainty estimates for Germany and its subregions. Derived quantities such as cumulative counts and 7-day incidences with corresponding uncertainties can be computed. The estimation of the time-dependent infection rate leads to an estimated reproduction factor that is oscillating around one. Data-driven estimation of the dark figure purely from incident cases is not feasible.

**Conclusions:** We successfully implemented a procedure to forecast near future COVID-19 incidences for diverse subregions in Germany which are made available to various decision makers via an interactive web application. Results of the incidence modeling are also used as a predictor for forecasting the need of intensive care units.

## 1 Background

Mathematical models of infectious disease epidemiology have experienced a boost of attention since the beginning of the COVID-19 pandemic. One can divide these models into three categories according to their purpose: scenario simulation, now-casting, and forecasting.

Scenario simulation focuses on different assumptions about some aspects of the model in order to compare and illustrate differences between several scenarios of in principle conceivable progressions of the transmission and other dynamics, which do not allow for proper uncertainty assessment. These approaches are used to examine the impact of changing certain parameters in the system, e.g. social behaviour, vaccination rate, etc. Nowcasting focuses on the precise description of the present situation based on incomplete, noisy and/or systematically biased data about the current state ([1], [2]). Forecasting tries to make predictions about the near future providing policy makers with reliable estimates of advancing developments. Similar to nowcasting, forecasting is strongly oriented towards realistic settings. The work presented in this publication focuses on a near-future prediction and can therefore be classified as forecasting.

### 1.1 The SPoCK Project

In Germany, local health authorities collect data about the infection dynamics on population level as mandated by the *Infektionsschutzgesetz* (IfSG) and report it to the national public health institute, the *Robert Koch-Institut* (RKI). In this paper, We describe the fitting and short term forecasting of this quantity, i.e. the newly reported cases of COVID-19 in Germany.

In a second step, which is not covered in this publication, the data about COVID-19-specific occupation of beds in intensive care units which is collected and reported daily by the *DIVI Intensivregister* run by RKI with support of the *Deutschen Interdisziplinären Vereinigung für Intensivund Notfallmedizin* (DIVI), is fitted and forecasted by our cooperation partners. The results of the first step are utilized as a predictor to obtain short-term future predictions on the level of intentsive care unit (ICU) capacities. This two-step procedure is referred to as the *Steuerungs-Prognose von Intensivmedizinischen COVID-19 Kapazitäten* (SPoCK) project. Several decision makers including the *Federal Ministry of Health* (BMG), the *Robert Koch Institute* (RKI), the *Federal Office of Civil Protection and Disaster Assistance* (BBK), the local planners of ICU capacities as well as the *Bundesamt für Bevölkerungsschutz und Katastrophenhilfe* (BBK) incorporate these predictions into their risk assessment of the current COVID-19 situation. In addition, the predicted incidences are visualized on an interactive web application provided by the *Deutsches Luft- und Raumfahrtzentrum* (DLR) called *Pandemic Mapping and Information System for Germany* (panDEmis).

The workflow within the SPoCK project is depicted in Figure 1. In this paper, we describe the daily analysis and prediction of incident cases of COVID-19 in different regions in Germany which are, in addition to the entire country, the 16 federal states (*Bundesländer*) and their 413 counties (*Land- und Stadtkreise*) summing to a total of 430 regions.

**Figure 1.**
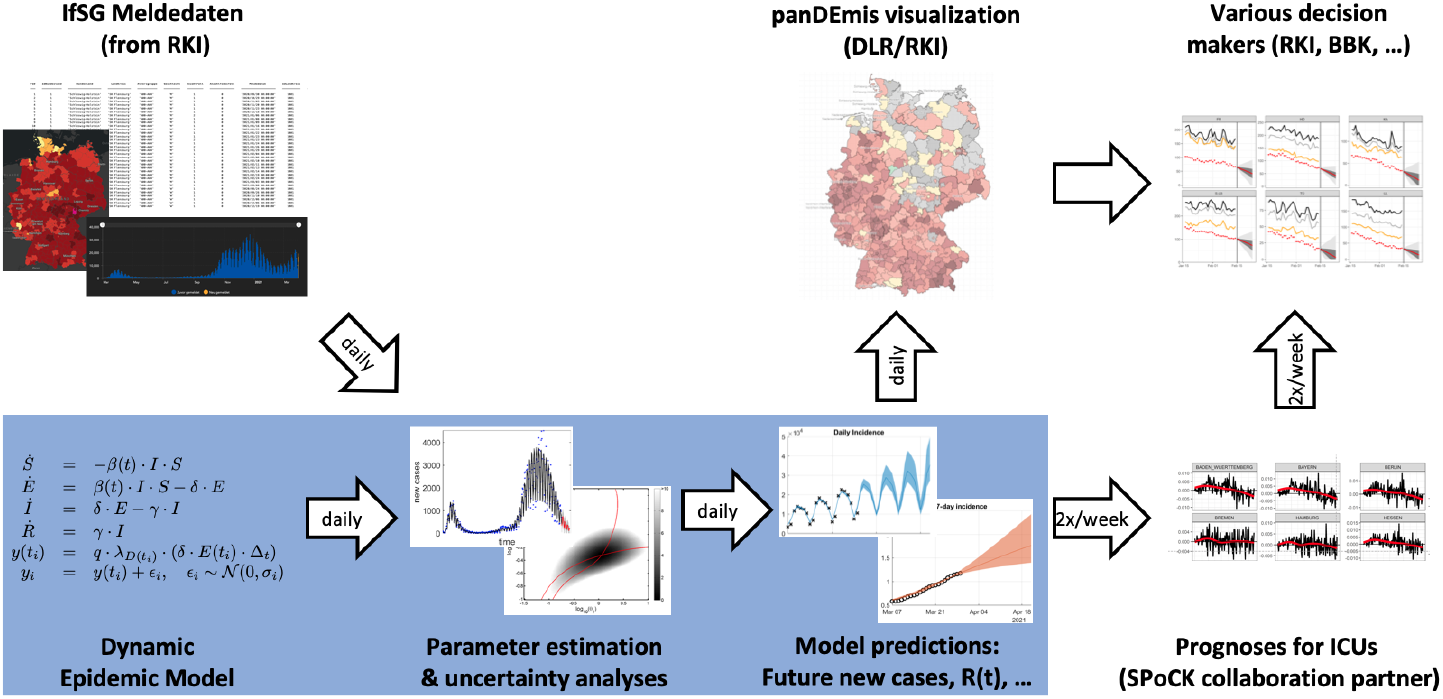
Schematic workflow of the SPoCK project. The SPoCK project predicts the needed hospital capacity of ICUs for COVID-19 patients. A key ingredient is the number of newly reported cases from the RKI which also has to be predicted (indicated by blue box). Results are used for visualization by the DLR and by decision makers, such as the BBK and RKI as well as local and regional health authorities.

## 2. Methods

A standard approach when describing infectious disease transmission are compartmental models or SIR-like models [3]. In general, both approaches divide the population into subpopulations with disjoint properties. Transition rates allow for flows between the subpopulations and define, in combination with the initial values of the subpopulations, the time evolution of the system. The ordinary differential equation (ODE) representation of the compartmental scheme we use is the well-known SEIR model [4]:

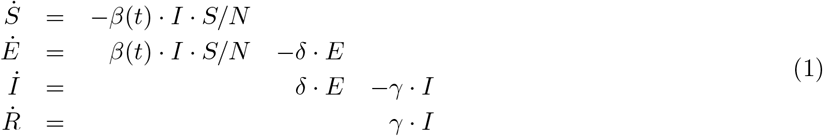

with *N* = *S* + *E* + *I* + *R* and where the dot notation is used to indicate time derivatives. A special characteristic of the current pandemic is the massive political and social reaction. In contrast to, e.g. the annual influenza season during which the social and professional life used to proceed pretty much as usual, the COVID-19 pandemic has led to vast political interventions and personal restrictions aiming mainly at the reduction of infections [5]. Within the SEIR scheme these changes over time can be described by a time-dependent infection rate *β*(*t*). There are several studies dealing with this problem in different manners. For example, at the beginning of the COVID-19 pandemic the impact of different non-pharmacological interventions (NPIs) was examined via step functions that implement *β*(*t*) via different variants of (smoothed) step functions, e.g. to examine the impact of different NPIs [6, 7, 8, 9]. Often, these approaches are restricted to time ranges in which the infection rate is assumed to be constant or monotonously decreasing or increasing, respectively.

In contrast, we aim for a more general approach which enables the infection rate to vary flexibly, i.e. to decrease and/or increase repeatedly within the considered time range. This is necessary for an accurate description of the COVID-19 transmission dynamics since it is influenced by many factors that may vary over the course of the ongoing COVID-19 pandemics:

1. Various NPIs are implemented, repealed and reintroduced iteratively.
2. The population’s compliance to regulative measures changes over time.
3. Seasonal effects, e.g. weather conditions, lead to changes in infection risk.
4. Mutations alter the physiological mechanisms underlying the disease transmission and other aspects.
5. Vaccinations reduce the population’s susceptible fraction.

In order to fit a strictly positive and time-dependent infection rate simultaneously with the SEIR model’s parameters, we introduce the following parametrization for the infection rate:

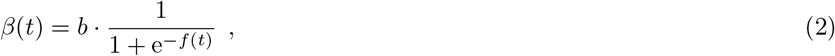

where the argument of the exponential function is given by an interpolating cubic spline

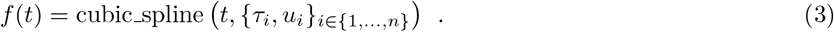

We utilize joint estimation of input spline and ODE parameters as introduced for biological systems in [10]. The composition of the interpolating spline (3) with the logistic function (2) allows for a nearly arbitrary time dependence, while still ensuring that the infection rate *β*(*t*) is strictly positive, smooth and restricted to a maximal value *b*. The cubic spline curve is determined by estimated parameters *u*_*i*_ = cubic spline(*τ*_*i*_) that represent its values at fixed and evenly spaced dates *τ*_*i*_ for *i* ∈ {1, …, *n* − 2} which cover the time range of observed data. In our model, the last two spline knots are placed after the date *t*_Last_ of the last data point: *τ*_*n*−1_ = *t*_Last_ + 50d and *τ*_*n*_ = *t*_Last_ + 300d. The value *u*_*n*−1_ is fitted to allow for some flexibility in the most recent regime, whereas *u*_*n*_ = 0 is fixed for numerical stability and reflecting the end of the pandemic in at least 300 days.

The predictions for the infection dynamics are primarily determined by the time-dependent infection rate *β*(*t*). In general, assumptions for the future development of *β*(*t*) are difficult to justify as many different factors contribute to it. For illustrative purposes, several different assumptions could be made and visualised as done e.g. in various online simulator tools [11]. For example, one such scenario study nicely illustrates the effectiveness of a Test-Trace-Isolate strategy [12].

For a data-driven approach focused on short-term forecasts, we need to be more practical: For extrapolation purposes, we fix

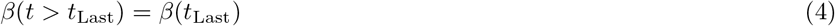

i.e. we assume the infection rate to be constant starting from the day where the last data point is reported.

### 2.1 Data-Driven Approach

Typically, there exist a multitude of model classes and structures which can be used to describe the same phenomenon. However, it is generally not possible to transfer results about estimated parameters between different models in a straightforward manner due to their differing mechanistic structures. To circumvent this problem, we here rely on a purely data-driven approach meaning that no prior knowledge about parameter values is incorporated into the optimization procedure. The only three *a priori* fixed parameters are the initial number of individuals in the susceptible, the exposed and the recovered state: *S*_init_, *E*_init_ and *R*_init_. Time point zero *t*_0_ is set to the first day that has at least a total of 100 reported cases to ensure the well-mixing assumption of ODE modeling. *S*_init_ was set to the total population of the respective region as given by the Federal Statistical Office of Germany [13]. *E*_init_ was set to *γ* · *I*_init_*/δ*, which is motivated by the assumption that *İ* ≈ 0 at the beginning of an epidemic reflecting a slow onset. *R*_init_ is set to zero. The only remaining initial occupation number *I*_init_ is estimated from the data.

### 2.2 Link between Model and Observed Data

In order to calibrate the ODE model, it needs to be linked to the observed data. The data we use for calibration is the daily incidence *y*_*i*_ published by the reporting date (*Meldedatum*) *t*_*i*_ at the local health authority. Therefore, we introduce the observation function

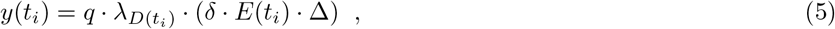

where the parameters canbe interpreted asfollows:

- *q* ∈ [0, 1] is the fraction of all infectious individuals that are detected and reported.
- *D*(*t*_*i*_) ∈ {1, …, 7} is an index for the weekday at date *t*_*i*_ where {1, …, 7} are naturally identified with the weekdays *W* = {Monday, …, Sunday}.
- *λ*_*D*_ is a factor for the weekday *D* that adjusts for the weekly modulation occurring in the IfSG data (see 2.2.1).
- (*δ* · *E*(*t*) · Δ) approximates the influx into the state *I*(*t*) of equation (1). As the considered data represents daily incidences, we set Δ to 1 day. This approximation of the true incidence quantity 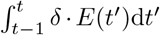 is exact if the state *E*(*t*) remains constant within that day. Comparison with this exact but computationally much more expensive approach showed minor deviations for real data applications.

The observable function (5) connects the model’s predictions to the reported data. The observations are assumed to scatter around this mean according to a normal distribution:

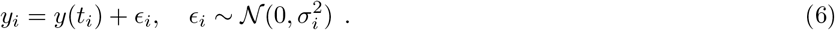

As we are dealing with a count process we use the standard deviation inspired by a Poisson model

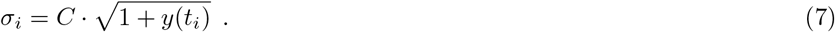

The error parameter *C* is fitted jointly with all others.

#### 2.2.1 Weekly Modulation Factors

The IfSG data shows an oscillatory pattern with a period of one week. The main reason for this is the reporting procedure, displaying a major delay during weekends, instead of actual infection dynamics. Therefore, we account for this effect within the observation function via seven weekday-specific factors *λ*_*D*_ with the integer *D* ∈ {1, …, 7}. In order to

1. guarantee that the factors *λ*_*D*_ essentially do not change the 7-day-incidence and
2. separate the weekly modulation from a global scaling of the observation function, which is realized via the factor *q*,

we, furthermore, set the constraint that

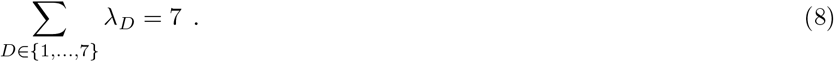

As a consequence, we are left with six degrees of freedom to describe the weekly effects. For a convenient implementation in the used software, we introduce a Fourier series with six parameters Θ_weekly_ = {*A*_1_, *A*_2_, *A*_3_, *ϕ*_1_, *ϕ*_2_, *ϕ*_3_}:

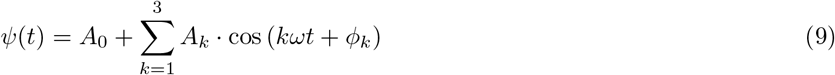

where offset and frequency are fixed to

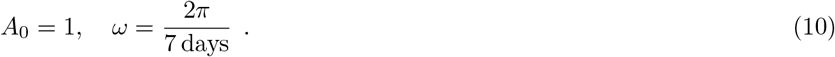

Instead of fitting the factors *λ*_*D*_ directly, we rewrite them in terms of equation (9) as

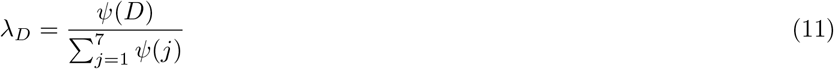

and calibrate the parameters Θ_weekly_. Doing so allows to set the amplitudes *A*_1_, *A*_2_ and *A*_3_ to zero in order to get an adjusted curve that does not feature the weekly oscillations and therefore reflects the ideal case of no reporting artifacts in the data.

#### 2.2.2 Correction of Last Data Points

The IfSG data published on date *t*_*n*_ contains information about the reported cases at all past dates *t*_*n*_, *t*_*n*−1_, …, *t*_1_ since the beginning of reporting. However, due to reporting delays between the test facilities, the local health authorities and the RKI, the data update from date *t*_*n*−1_ to *t*_*n*_ contains not only cases that were reported to the local health authorities at date *t*_*n*−1_, but also before that at dates *t*_*n*−2_, *t*_*n*−3_, … and so on. This means that the number of reported cases on day *t*_*n*_ will be underestimated especially for the most recent dates. For some regions this correction factor be as big as three for the most recent day.

Meaningful handling of this data artifact can be done in at least two ways: For instance, one could choose to ignore some of the latest data points, since they are most prominently affected by this data artifact. An alternative is to estimate the systematic deviation from historically published data sets. In order to avoid the bias towards smaller incidences in the prediction, the data can be adjusted accordingly. Therefore, one assumes, that the future data sets of *t*_*n*_ will not change reported counts older than four weeks *t*_*n*−28_. Let 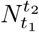 denote the number of reported cases, that were published at time point *t*_1_ to be reportedly infected at date *t*_2_ where 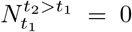 as future cases cannot be reported. Then, one can learn from this history of published data sets the correction factor *CF*_*k*_

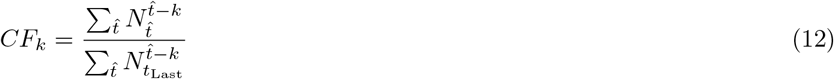

the initial publication of *k* day old counts had to be corrected to obtain the number in the latest data set *t*_*n*_. The factors *CF*_*k*_ can then be applied to the newest data set.

This was done for Germany and all the federal states separately. We showcase the resulting differences of these two data preprocessing strategies in section 2.4.2.

For the county level, this adjustment is not as crucial for two reasons: 1) the count numbers are much lower, so the stochasticity can lead to wrong correction factors and 2) the shape of the estimated dynamics is inherited from the federal states in our model.

### 2.3 Parameter Estimation

In general, we follow the maximum likelihood estimation (MLE) approach. As there are a total of 429 regions for which the data has to be fitted and predictions are calculated, we rely on a two-step procedure to reduce computation time which is described in the following paragraphs.

#### 2.3.1 Federal States and Germany

The parameter estimation problem given by the above defined ODE model and the IfSG daily incidence data is solved separately for Germany and each federal state by an MLE approach. The latter has been well established for ODE models [14]. The deviation between data and the model’s observation function as specified in equation (5) is minimized, taking into account the error model of equations (6) and (7). The simultaneous parameter estimation of the spline parameters *u*_*i*_ follows the lines of [10]. In particular, no explicit regularization term is implemented that penalizes non-vanishing spline curvatures.

#### 2.3.2 County Level

Analysis at the rural and urban county level (*Land- and Stadtkreise*) is important to obtain a spatially resolved picture of the infection dynamics in Germany. The previously described approach is computationally not feasible because the analysis of 429 regions cannot be performed within 24 hours without access to a sufficiently large computing cluster which can be used 24/7 without queuing. Moreover, the number of infected individuals can generally be so small at the county level that inference and prediction based on a purely deterministic model is not appropriate. Therefore, we used the results on the higher-level administrative structure, i.e. the fitted model of the federal state, as prior information about the dynamics, and scaled it down to the county level for predictions.

More specifically, the county-level data was used to merely estimate two parameters in a county-specific manner: the scaling parameter *q* from equation (5), which in this context can be related to the proportion of current infections occurring in the county *c*, and the error parameter *C* from equation (7) which quantifies the stochasticity of county-level observations analogous to its meaning on the level of federal states. All other parameter values for a county *c* are taken from the estimated set of parameters 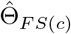 for the corresponding federal state *FS*(*c*).

The county-level dynamics might change rapidly as new clusters of infection emerge. For predictions, it is important that such rapid changes are detected by the model calibration procedure, i.e. fitting of *q* and *C* has to account for such rapid changes. We implemented this requirement by exponentially weighting down the county level data observed in the past by increasing the standard deviations via

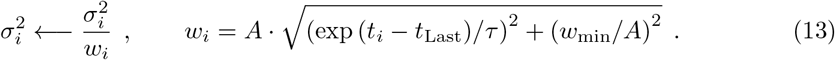

Here, *w*_min_ = 0.01 · *A* denotes the minimal weight factor used for data observed in the past. *A* = 7.56 denotes the normalization factor that ensures that the sum of all weights *w*_*i*_ is equal to one. Moreover, we chose *τ* = 7 as time-constant of this weighting step. To be clear, on the county-level, *σ*_*i*_ from equation (7) should be thought of as first being transformed according to the mapping (13) before entering equation (6) as the standard deviation of Gaussian observation errors.

Just as the analysis for the federal states, the described scaling procedure for the counties is updated on a daily basis, i.e. the county-specific parameters *q* and *C* are updated every day. This accounts for time-dependent deviations of the local infection history on the federal state level, i.e. each county has an individual kinetics.

### 2.4 Calculation of Uncertainties

To quantify the uncertainty in the predictions of the model, our forecasting tool provides confidence intervals along with proposed predictions. Here, we describe two main sources of uncertainties: parameter uncertainty and approach uncertainty. The first is captured by simulating all parameter combinations that agree with the observed data as will be explained in section 2.4.1, the second is incorporated by running the analysis with several models as detailed in section 2.4.2.

#### 2.4.1 Profile Likelihood Analysis

For non-linear models, uncertainties for estimated parameters can be determined using the *profile likelihood* (PL) method which estimates parameter values that still ensure agreement of model and data to a certain confidence level in a pointwise and iterative manner [15]. This approach has been showcased for infectious disease models [16]. Parameter uncertainties naturally translate to prediction uncertainties which can be analyzed systematically [17]. Following the given references, we simulate the data-compatible parameter combinations from the parameter profiles and then take the envelope of the resulting family of curves to obtain confidence intervals.

One could also analyze the uncertainty of a model prediction directly via the *prediction profile likelihood* method [18]. Prediction profiles need to be computed via a costly iterative fitting procedure for each predicted quantity and time point separately. However, by using the parameter combinations from the profile likelihood method, we can calculate uncertainties for any desired model quantities and time points only by simulation, thus rendering this method more efficient for our purposes.

#### 2.4.2 Averaging of Approaches

When utilizing ODE models to describe certain aspects of reality, a multitude of assumptions are implicitly made, which include (but are not limited to) the selected model structure, the noise model of the data, the appropriate data preprocessing. All these decisions result in a certain *approach*. These necessary decisions along the modeling process impact the space of possibly described and therefore also predicted dynamics. To account for this origin of uncertainty, we perform the procedure described so far simultaneously for several approaches and merge their results into one comprehensive result. The latter is done by taking the mean / minimum / maximum of the different approaches’ MLE / lower bound / upper bound curves. Accounting for different modeling decisions prevents overconfidence in the results.

## 3 Results

Since April 2020, the described methodology has delivered daily predictions and the *ansatz* has evolved and several changes and refinements have been implemented. Currently, the resulting predictions for ICU bed capacity, which use the here presented results of estimated incidences as a main predictor, are reported bi-weekly to public health decision makers. The presented methodology and results were generated on April 1st, 2021. The data fitted had therefore registered infections up to March 31st, 2021.

### 3.1 COVID-19 Spread in Germany

For the aggregated data over all of Germany, we obtained a fit and predictions with uncertainties as shown in Figure 2. The data can be described by the model. Adjusting for weekday effects turned out to be beneficial and the prediction is a reasonable continuation of the last data points. The most interesting model quantity is the time-dependent infection rate *β*(*t*) which translates to an effective time-dependent reproduction number 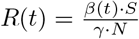. The latter quantifies how many other people are infected on average by a single infectious individual and determines at which rate the number of currently infectious individuals is growing (*R*(*t*) *>* 1) or decaying (*R*(*t*) *<* 1). It should be noted that, despite the fact that *β*(*t*) is extrapolated as remaining constant (see equation (4)), *R*(*t*) is not necessarily constant. This is because *R*(*t*) includes the monotonously decreasing susceptible density 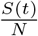.

**Figure 2.**
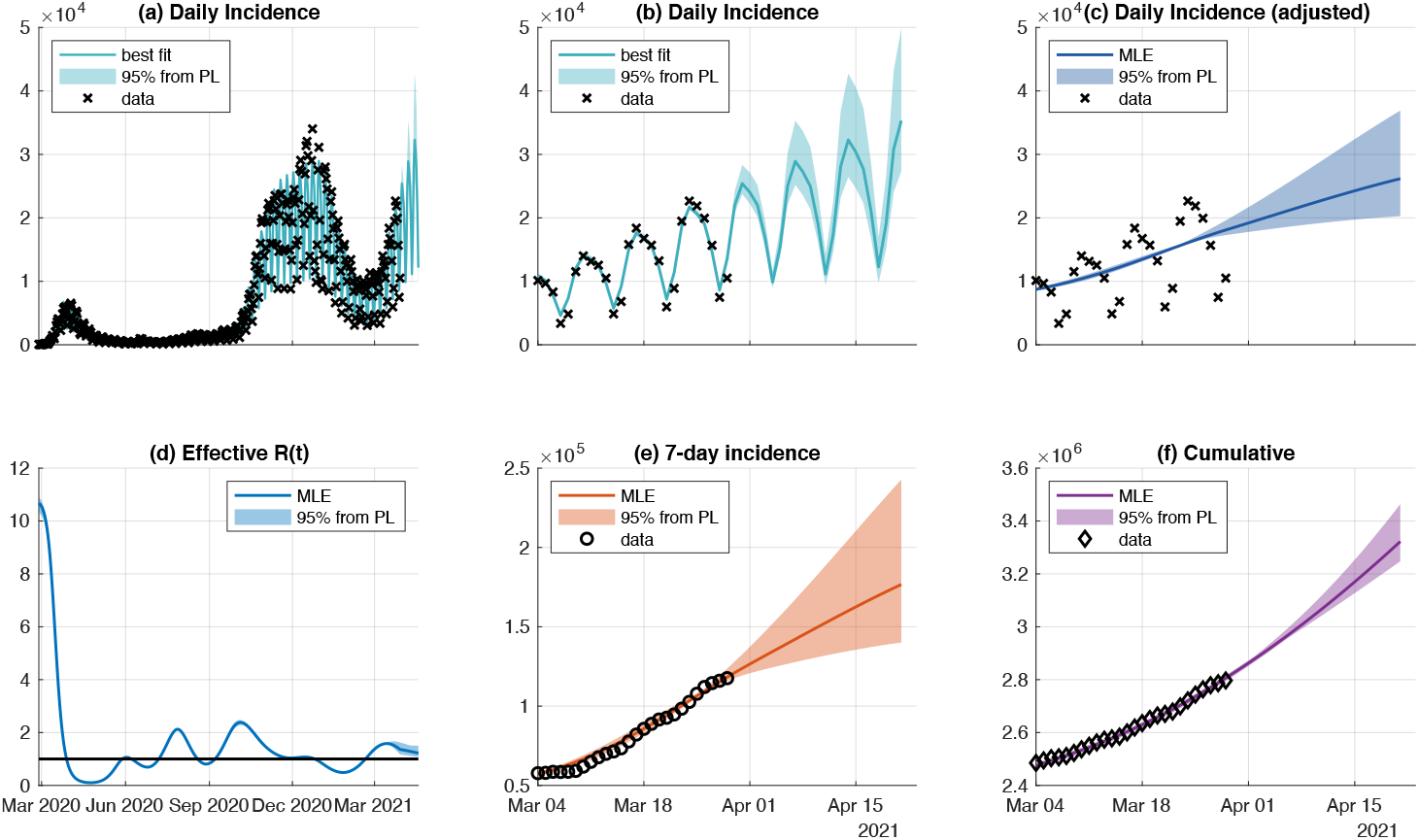
Fit and prediction for Germany. The incidence data of the entire time course is fitted (panel a) to estimate all dynamic parameters including the time-dependent infection rate that corresponds to *R*(*t*) (panel d). Predictions of incidences (panels b and c) and derived quantities (panels e and f) for a zoomed in time span are shown. 95%-confidence intervals (color-shaded areas) are inferred by profile likelihood calculation.

The estimated reproduction number *R*(*t*) oscillates around a value of 1 and illustrates the effect of describing the politics’ countermeasures and the population’s compliance to them (Figure 2, Panel (d)). This is in line with several publications [9], [19], [20] reporting similar behavior of the reproduction number. In general, oscillations in dynamical systems often are attributed to a feedback with delay, which is also the case here for the reproduction number *R*(*t*). Several additional quantities of interest, such as the 7-day incidence or the cumulative number of cases can be computed from the model’s predictions. In addition, the associated confidence intervals of these quantities can be determined using the parameter sets below the 95% threshold of likelihood profiles. We stress here again, that only the incidence data was used for model calibration (Figure 2, Panel a).

### 3.2 COVID-19 Spread in Subregions of Germany

For the county-level (*Landkreise*) we obtain results by the scaling approach described in section 2.3.2. The shape of dynamics is preserved and describes the latest data. Due the exponential scaling on later data points, it is unlikely that the entire time course is described well by the scaled dynamics. As we are primarily interested in the forecast, we display only the latest time interval. The data is more noisy due lower numbers of cases and inhabitants (Figure 3). Here, we show already merged results for clarity (see section 3.3).

**Figure 3.**
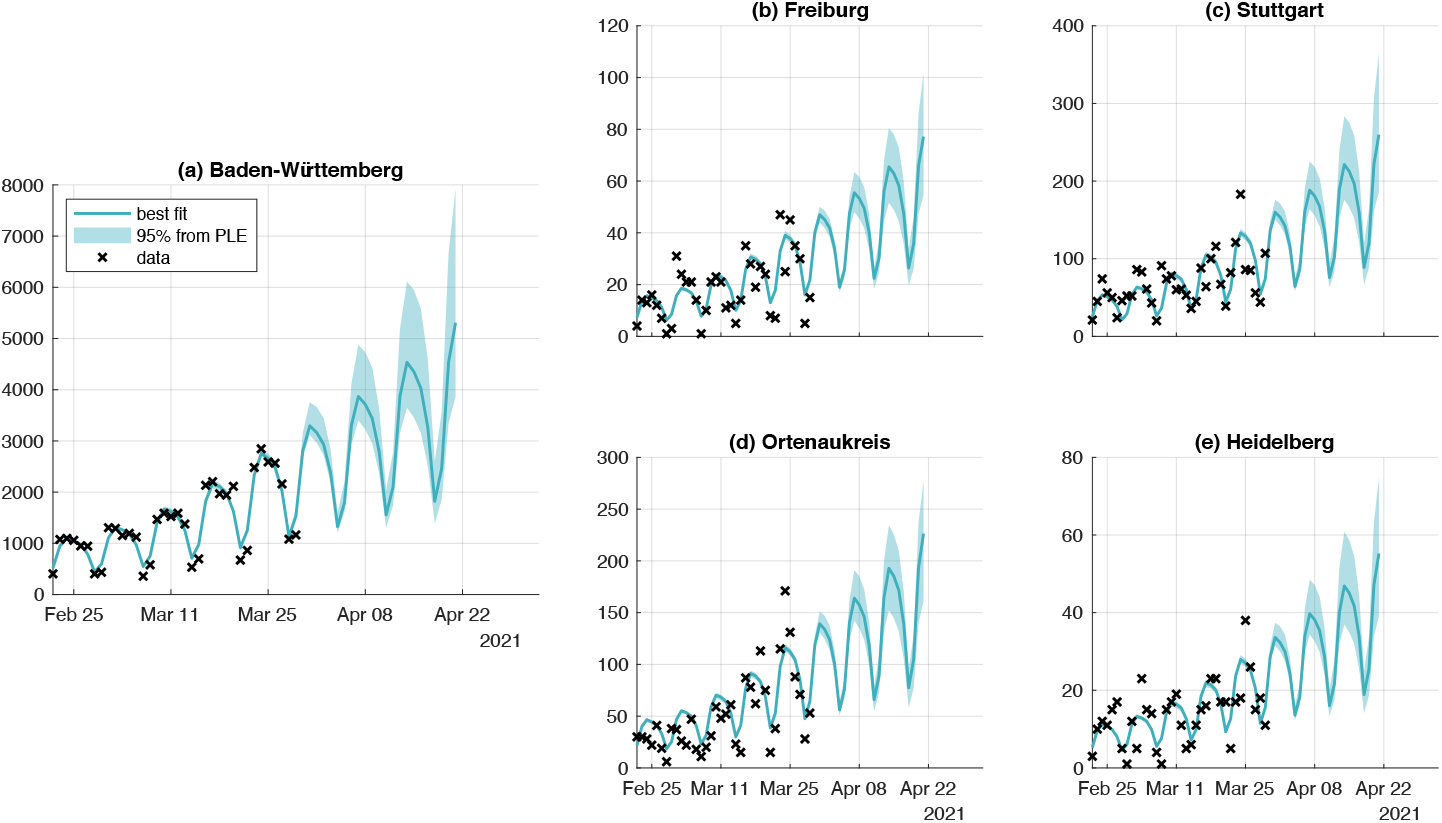
Fit and prediction for one Bundesland and four counties. The dynamic of the one exemplary Bundesland Baden-Württemberg (panel a) governs the dynamics of the corresponding Landkreise, four of them are shown here (panels b through e). For regions with fewer inhabitants, lower case numbers are expected: note the different scaling of the y-axis for Bundesland and Landkreise.

### 3.3 Approach Averaging

The analyses can be carried out for different approaches representing a variety of *a priori* equally feasible modeling strategies. To account for the uncertainty that arises from (possibly over-)simplifying modelling assumptions, those different approaches are analyzed independent from each other. After results for all regional entities, i.e. federal states (as in 2 and counties 3) have been obtained for each approach, the results are merged into one comprehensive prediction, which features by construction (see 2.4.2) a higher uncertainty, now including both the uncertainty in the data and the uncertainty which modeling strategy is used. We illustrate this for two different approaches which differ only in the handling of the most recent data points (Figure 4). In general, this methodology generalizes to an arbitrary number of different approaches with the available computing resources as the only limiting factor.

**Figure 4.**
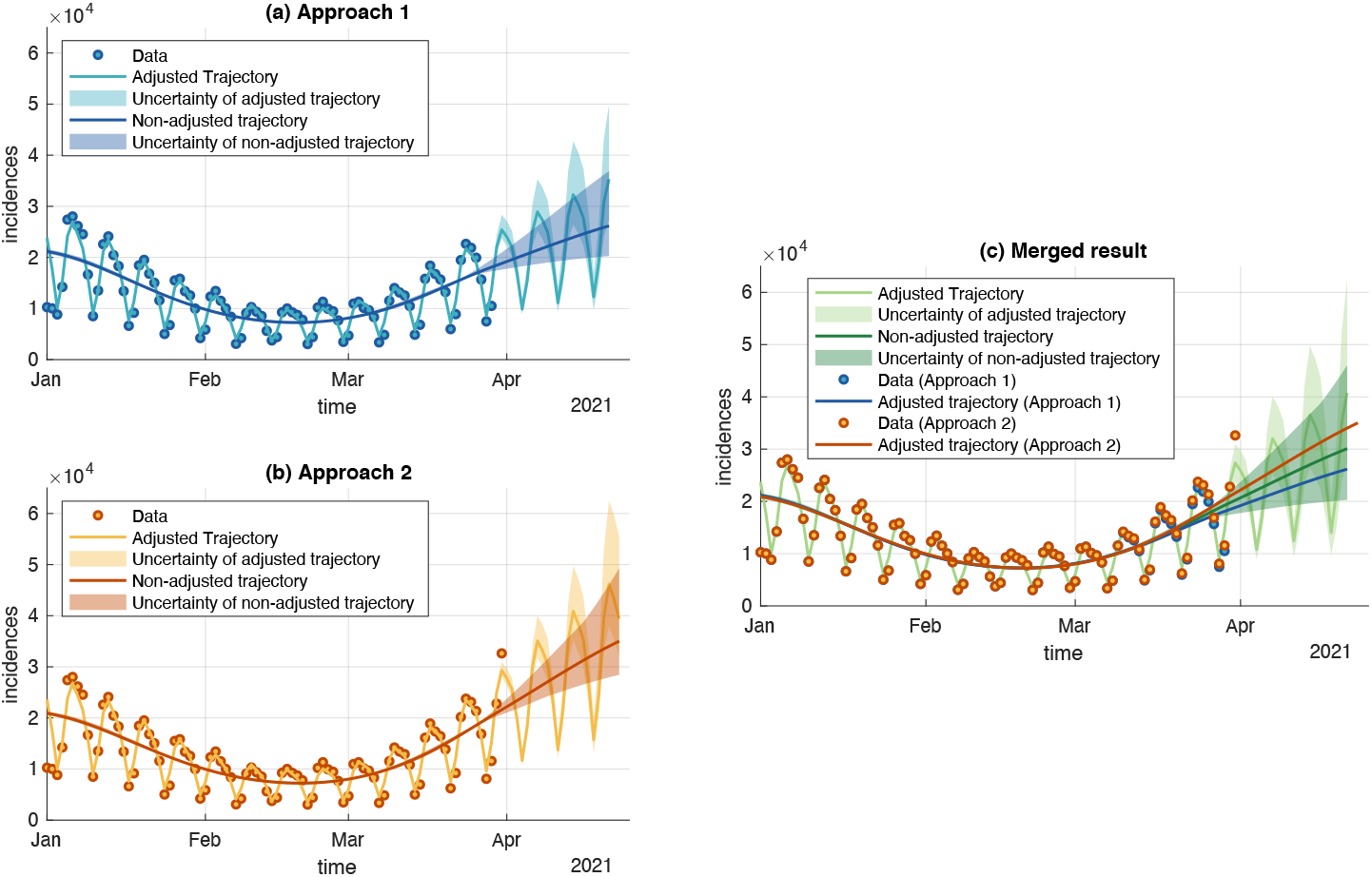
Merged Approaches for the example of Germany. The two approaches differ in their data handling strategies for considering reporting delays: Approach 1 (panel a) simply ignores the two latest data points. Approach 2, in contrast, uses estimated correction factors on the latest data points (panel b). The result of the merging (panel c) indicates that both approaches describe the data well, but make differing predictions. Therefore the resulting uncertainty is bigger than the individual uncertainties. In general, this procedure generalizes to more different approaches.

### 3.4 Availability of Results

Sound political or social decisions are based on an empirical or prognostic foundation. To make the daily generated predictions available to various stakeholders, the forecasts are integrated into a web-application called panDEmis: In this interactive application, the recent infection situation is analyzed and displayed. For all registered users of the *DIVI Intensivregister* the tool is available at https://pandemis.dlr.de/de/#/overview. Current capacities of hospital beds and intensive care units, exposed population in the catchment areas of hospitals are merged with the forecast data. The combined display of all available data sets allows a situation picture for each day including also for past and future time steps. Figure 5 shows different features of the web-application from May 17th, 2021 for the occurrence of infection in the map entire Germany (panel b), as well as for the selected administrative district of Bayern (panel a). Here, the blue graphs represent 1) the daily reported new infections by RKI, 2) the incidence of COVID-19 cases in the past 7 days per 100,000 people and 3) the cumulative infections. The prognosis is displayed as red curve, including a 95% confidence interval. All data can be interactively analyzed and visualized for different administrative units, i.e. federal states and county level.

**Figure 5.**
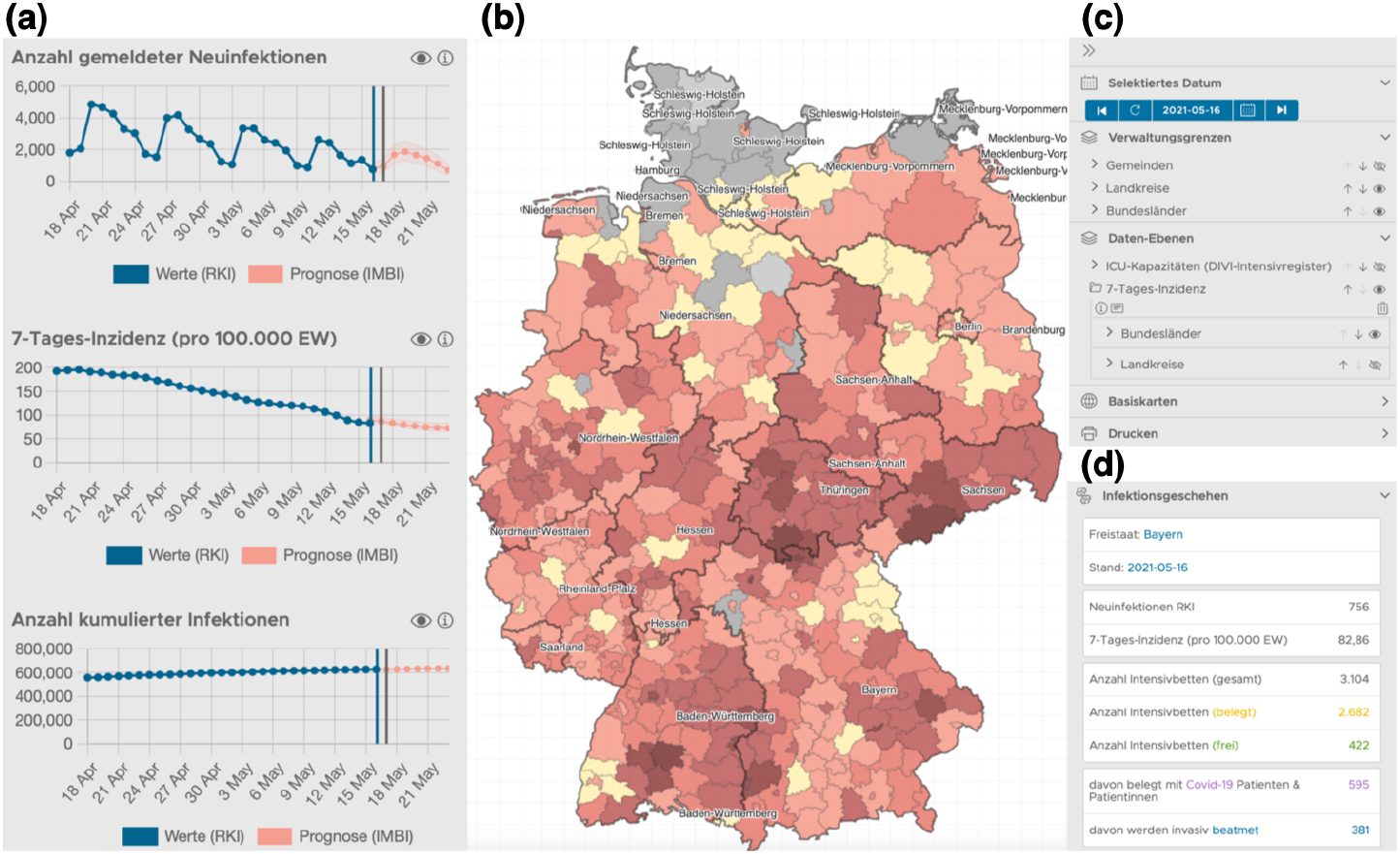
panDEmis visualization. On the interactive web application called panDEmis, predictions for incidences, 7-day average, as well as cumulative cases can be inspected for all subregions (panel a). The region can be selected through a map indicating all the regions (panel b). For the chosen regional district, historic data sets and predictions can be selected and different layers can be chosen for visualization (panel c). Additionally, key figures about the current pandemic situation, such as incidences and ICU bed capacities are displayed for the selected region (panel d).

The results of this incidence modeling approach are also a main predictor for a prediction analysis of ICU beds. The results of this second analysis step which is not detailed within this paper, is available for all registered users of the *DIVI Intensivregister* at https://www.intensivregister.de/#/aktuelle-lage/prognosen.

## 4. Discussion

Different model classes as ODE models or stochastic differential equation (SDE) models with or without mixed effects could be used for a data-driven parameter estimation approach. An SDE approach might be beneficial for small regions with low infection numbers or during times with very low total infection numbers. In these cases local outbreaks dominate the infection dynamics and the population is not *well-mixed* which renders an ODE approach ineffective. For the presented regional entities, the underlying assumptions for ODE modeling are reasonable and the ODE model was successfully adapted. We here focused on a pragmatic procedure that allows daily analysis and reliably calculates predictions.

When fitting data about the number of reported cases of an infectious disease outbreak, it is beneficial to fit incidences (or fluxes) instead of the total (or cumulative) number of cases [21]. The residuals of a fit on cumulative data will be correlated by construction. Most noise models assume independent measurement errors. Thus, the uncertainty will be underestimated in these cases and obtained results will be overly confident. By fitting the model to incidence data, the measurement errors remain uncorrelated.

The presented modeling approach heavily relies on the time-dependent infection rate *β*(*t*). We assume dynamic processes to be continuously differentiable which leads to a smoothing of possible steps in the real infection rate which might occur due to rapid policy changes. Also, the temporal change *β*(*t*) incorporates many different mechanisms, which include but are not limited to: vaccinations, NPIs, changes in compliance to NPIs, viral mutations, seasonality and testing frequency. For an assumed constant vaccination rate, we saw that our approach delivers the same results when omitting the explicit vaccination state since *β*(*t*) is flexible enough to compensate the vaccination effect.

In general, it is *a priori* unclear how much flexibility this function should have. In the presented procedure, this corresponds to the number of knots employed in the spline. The spline’s freedom should allow for a good fit of the dynamics, but also prevent overfitting.

Furthermore, the dynamics of the prediction are primarily determined by the value of *R*(*t*) at the latest data point. Hence, this value should not be estimated by too few data points meaning that the last spline knot should not be too close to the end of the time series.

Any prediction model used for forecasting should not exceed a certain time period as the future infection rate is hard to determine. But even at a short prediction time span, it is unclear how recent political measures and the population’s resulting behavior will alter the future infection rate. Therefore, we assume *β*(*t*) to be constant starting at the last data points. By additional precise knowledge about the effect of planned or recently made political decisions or other effects like weather conditions, this assumption could be further refined.

In contrast to other modeling approaches, we do not feed the actual NPIs into the model, but can instead correlate the estimated time development in a second step of the infection rate to NPIs. Quantifying the NPIs’ effect and time lag on *R*(*t*) is difficult as most NPIs are not imposed or lifted independently of each other and estimates will therefore be highly correlated [22].

Whenever discussing the required amount of flexibility to obtain a good model fit, one should be aware of bias-variance-tradeoff: The introduction of more parameters included to explain a certain time dependence (reducing the bias), the bigger the resulting prediction uncertainty will be (increasing the variance). Similar arguments can be made when discussing the amount of utilized spline parameters or accounting for age structure. More available and consistent data can help.

There are no explicit states in our model to distinguish between recovered and dead people, mainly for the reason that there is no reliable data over the entire time course for those quantities. Recovered individuals are not tested to be non-sick anymore, and people who died were not consistently assessed in real-time in Germany.

Furthermore, the unobserved infected and infectious individuals are not in an explicit state. This fact is compensated by two aspects: Firstly, the used data does not contain information about the duration from beginning of infectivity to reporting to the local health authority. Thus, since the additional state would not help to better describe the used data, it is omitted. Secondly, the factor *q* introduced in the observation function in section 2.2 accounts for individuals that are overseen at all times. The estimated dark figure from equation (5) when fitting only incidence data is in the presented modeling approach in most regions compatible with a broad set of values ranging from 0.1 to 1 within the confidence level. This means that anywhere between 10% to 100% of all cases are detected by local authorities and both edge cases still agree sufficiently with the data. Therefore, the dark figure can not be estimated solely based on reported incidence cases. For reliable determination of the dark figure, additional testing in pre-specified cohorts is necessary.

## 5 Conclusions

We presented a data-driven ODE approach to fit and predict incidences of COVID-19 cases for different subregions of Germany. The key ingredients in doing so are 1) likelihood-based estimation and uncertainty quantification and 2) a time-dependent infection rate which is estimated by utilizing a cubic spline. All parameters are estimated from data and uncertainty in parameter estimates are translated to prediction uncertainty. As many different modeling assumptions will affect the outcomes, we average over similarly plausible approaches to account for this source of uncertainty. A major constraint for a feasible analysis strategy is a maximum runtime of 24 hours as the analysis should be repeated on a daily basis in an automated manner including the respectively newest data set.

In the future, more work for validation of competing modeling approaches and comparison of the various efforts undertaken in the currently highly dynamic field of mathematical modeling of infectious diseases is needed and will certainly be seen.

## Data Availability

Public data was used to conduct the presented analyses.
Code for analyses and/or access to respective github repository can be requested via email.

## Competing interests

All authors have completed the ICMJE uniform disclosure form at www.icmje.org/coi_disclosure.pdf and declare: no support from any organization for the submitted work; no financial relationships with any organizations that might have an interest in the submitted work in the previous three years; no other relationships or activities that could appear to have influenced the submitted work.

## Author’s contributions

Lead conception of the overall study design: LG, HB; Conceived and designed the analysis: LR, FL, CK; Collected the data: MF, RKI; Contributed data or analysis tools: LR, FL, CK; Performed the analysis: LR, FL, CK; Integration and description of information system components (panDEmis): HT, TR; Wrote the paper: LR, FL, CK.

## Acknowledgements

The project was funded by the Bundesministerium für Gesundheit (BMG).

We thank Matthäus Lottes, Janina Esins and the team from DIVI Intensivregister at the RKI.

Also, we thank the RKI’s statisticians responsible for processing of the raw and routine data.

We thank Mario Menk, Steffen Weber-Carstens, Christian Karagiannidis, Uwe Janssens for fruitful discussions during project planning and implementation.

Thanks to Rafael Aruntjunjan for critically revising the manuscript.

## Notes

### Competing Interest Statement

The authors have declared no competing interest.

### Funding Statement

The project was funded by the the German federal ministry of health Bundesministerium für Gesundheit (BMG).

### Author Declarations

None to be reported.

